# Prolonged time to breast cancer surgery and the risk for metastasis

**DOI:** 10.1101/2024.01.11.24301150

**Authors:** Dieter Hölzel, Anne Schlesinger-Raab, Gabriele Schubert-Fritschle, Kathrin Halfter

## Abstract

**Background:** A growing breast cancer (BC) is associated with an inherent risk of metastasis (MET). If surgical treatment of a BC is delayed, the prognosis worsens continuously with increasing tumor diameter (TD). This frequently overlooked topic in healthcare is currently the subject of debate.

**Methods:** Population-based data on BC incidence and surgery waiting period for the U.S. and Germany are used to examine the resulting risks. BC growth and initiation of MET are calculated in a simulation approach using hormone receptor status (HR), tumor volume doubling time (VDT), and TD-dependent survival.

**Results:** The U.S. and Germany report 287,850/71,375 BCs annually. Based on an initial mean TD of 19.8mm 15-year mortality in both countries is estimated using a Gompertz function at 19.6% without surgical delay. VDT of HR+ and HR- BCs differs by a factor of 2.4, leading to an estimated mortality increase of 1.34/3.26% (HR+/HR-) if all surgical procedures are pushed back by five weeks. The mean delay in the U.S. and Germany is 33.7/26.0 days respectively. Prolonged waiting periods lead to estimated 4,676/918 additional BC deaths or a 1.6/1.2% rise in BC mortality rate. Further increases are due to MET relapse up to 25 years after diagnosis.

**Conclusions:** A growing tumor can continuously initiate MET with every millimeter. The results of this study offer replicable and valid evidence, based on publicly available clinical and population-based data, that confirmed BCs should be treated according to HR status as soon as possible.

## Introduction

The question of whether a prolonged time between diagnosis and the start of therapy poses a risk for cancer patients has been examined many times, most recently because of interruptions during the COVID-19 pandemic.[1] Based on the lessons learned from the pandemic a similar systematic disruption of health care provision in the future is unlikely. Yet looking at data from the U.S. and Germany the time between diagnosis and primary treatment continues to increase, especially in breast cancer (BC). Currently an eight week time period between diagnosis and surgical intervention is proposed as an upper limit in BC.[2] However, previous research such as a meta-analysis by Hanna et al found that a four-week delay across all common forms of treatment is associated with an increase in mortality in seven cancers.[3] The aim of this simulation study is to quantify and compare the association between a surgical treatment delay and mortality for the female BC populations from the U.S. and Germany.

## Methods

The time from the confirmed diagnosis of an invasive BC to the start of treatment is referred to as the delay and includes the time required for organizational and technical processes. The relationship between a prolonged waiting period, metastasis (MET), and mortality can be quantified based on publicly available data:

1. 15-year BC specific mortality is dependent on tumor diameter (TD) (Figure 1A). For locoregional BCs (T-N-M0), the relationship can be described with a Gompertz function based on population-based data from the Munich Cancer Registry (MCR).[4] 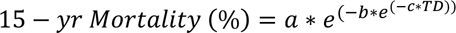 (with a: 58.4, b: 4.46, c: 0.071). Mortality thus increases by 6.8%, for example, with tumor growth from 12.5 to 17.5 mm or by 1.36% per mm.[5]
2. Volume doubling time (VDT) of BCs depends on hormone receptor status (HR). The mean values derived from two publications are 72/170 days for HR+ and HR- BCs. [6, 7]
3. Time to MET also depends on HR status. According to data from the MCR the 80% percentile intervals PI_80_(p_10_, p_50_, p_90_) of time to MET in M0 BC are PI**_80_**(16.9,60.4,141.3) months for HR+ and PI80(9.4,25.2,80.8) months for HR- (Figure 1D).[8] This implies that 50% of developing METs in M0/HR+ BC will be diagnosed within 5.0 years, and in M0/HR- BC within 2.1 years after primary treatment. If MET risk increases with delay, then these METs are initiated during this prolonged waiting period. This means that these METs become detectable after twice the MET-free time of 60.4/25.2 months on average. This is equivalent to the mean growth time of MET.
4. The distribution of HR status is important for quantification. Current data states that 85% of BC are HR positive.[4 9]
5. Population data on BC incidence are required for estimation. For the U.S., 287,850 newly diagnosed invasive BC are estimated in 2022, for Germany 71,375 BCs in 2019.[9],[10]
6. The distribution of delay to BC surgery (in weeks) is available for the U.S. and Germany (see table 2).[2], [11]

**Figure 1:**
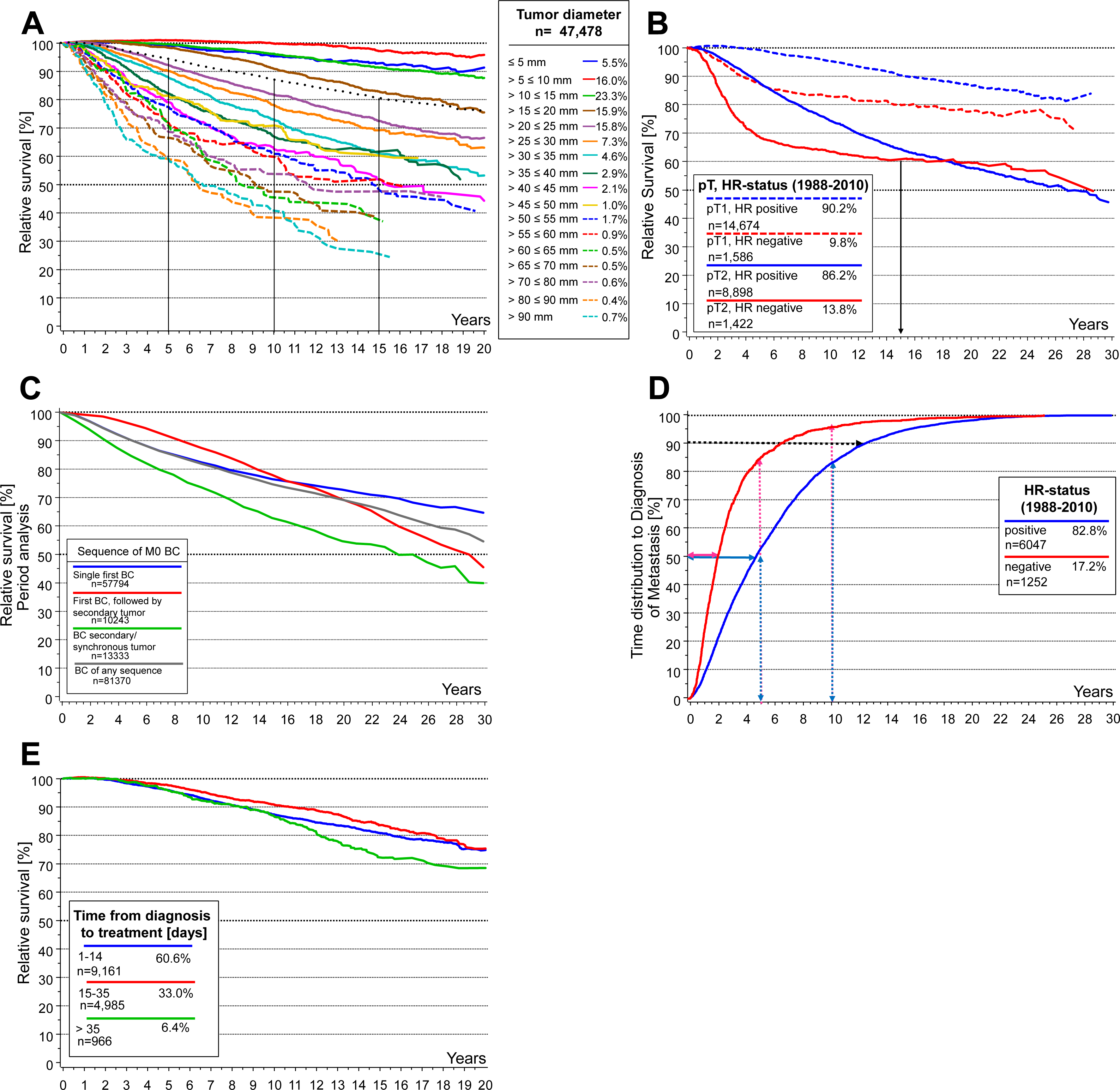
Long-term breast cancer survival based on data from the MCR. **A:** Relative survival in dependence on TD without neoadjuvant treated BCs. The dotted line shows the relative survival of all T-N-M0 BCs of 80.7% at 15 years and 76% at 20 years (1998-2019, n=47,478). **B:** Relative survival in dependence on hormone receptor status for first single BC (1988-2010, pT1 n=16,260, pT2 n=10,320). The faster growth of HR negative MET causes very different results in the pT1- and pT2-BC subgroups. **C:** Relative survival according to tumor sequence (Period analysis, BC M0, 1988-2018), only first single BCs adequately illustrate the BC-specific disease course, the other disease courses are impacted by preceding or secondary cancer diseases. **D:** Time to diagnosis of MET in dependence on hormone receptor status for first single M0 BC (diagnosed 1988-2010, n=7,299). Early MET are initiated long before diagnosis, late MET shortly before. Therefore, twice the median time is the median growth time of MET initiated during delay until detection. The growth time depends on tumor biology and not on the size of the BC at the time of MET initiating tumor cell spread. Secondary tumors are excluded because MET from a contralateral BC, for example, would prolong the time distribution. **E:** Relative Survival of hormone receptor positive and M0 BC in dependence on days delayed (1998-2010, n=15,112).

The relative survival is calculated from the observed lifelong all-cause survival within the cancer population and the respective expected all-cause survival of the general population using the official life table. The quotient of current mortality and incidence corresponds to the prognosis of the disease. It can be considered equivalent to the cure rate when the relative survival approximates this value and reaches a plateau (Fig.1B). Only very few MET still occur after 20 years. Interim advances in treatment over time can be adjusted for by using period survival analysis for prognosis.[12] The lifelong risk of secondary malignancies must be analyzed in the cure rate for long disease courses due on the basis of the cause of death.

In order to quantify the risk of MET and implications on survival through delay, we built a function based on TD for the relevant group of T-N-M0 BCs using MCR population-based survival data. The mean value of TD is 19.8 mm, which is associated with a mortality of 19.57% (± 0.5%) after 15 years at risk (Figure 1A). The prognosis is more favorable due to the exclusion of 6.2% primary M1 BCs from a prolonged waiting period to primary treatment. HR status is another important growth factor. Figure 1B depicts resulting short- and long-term survival according to HR for patients without secondary cancer. This leads to a more refined approximation of the BC-specific disease course with the times to MET as a function of HR status (Figure 1D). Second malignancy risk in BC, in particular the contralateral BC depending on event sequence is shown in Figure 1C.

### Statistical analysis

The presented data are used to estimate the effects of a prolonged treatment waiting period on BC mortality. Relative survival was calculated using the ratio of the observed to the expected survival rate. The latter was estimated using the Ederer II method and age- and sex-matched life tables of the corresponding population. Time to MET was estimated on the basis of MCR data by cumulative incidence analysis considering competing risks (e.g. death).[13] Multivariate regression and survival analysis were done with SAS V 9.4. For the simulation and the delay calculations, the R system version 3.1.3 was used.[14] For all analyses, a two-sided p value of 0.05 or less was considered statistically significant. Because of a legal change in cancer registry proceedings, MCR high quality follow-up data are only available until the 2018 patient cohort (censored on March 31, 2019).

### Ethical considerations

Only preexisting retrospective data with no personal identifiers was used in the study making it exempt from review and approval by an ethics committee or competent authority.

## Results

To estimate the delay risk, we calculated the growth of BCs as a function of HR status using a mean of two published values (72/170 days for HR-/HR+). Table 1 shows the growth depending on the delay in a HR+ and HR- BC with a TD of 19.8 mm. For example, a HR+ BC reaches a TD of 20.68 mm (19.8* 1.26 ^(32/170)^) within five weeks or up to day 32. A HR- BC would have grown to a TD of 21.94 mm, assuming the same initial TD of 19.8 mm. Applying the Gompertz function for 15-year mortality results in a mortality of 20.91%/22.83% for HR+/HR- after a surgical treatment delay of five weeks. In relation to the base value for a 15-year mortality of 19.57% without delay this corresponds to an increase of 6.8/16.7% for HR+/HR- BCs respectively (Table 1). Considering 1,000 BCs treated without delay, 196 BC deaths are expected after 15 years, a delay of five weeks would result in an estimated 13/33 additional delay-associated deaths for HR+/HR- BCs.

**Table 1:** Increase TD and 15-years mortality risk as a function of HR-status and delayed surgery in weeks. A TD of 19.8 presents the mean of the population-based T-N-M0 findings in Figure 1A.

| Delay<br>[weeks/<br>months]<br>Aspects:<br>cohorts n=1,000 | Basis | 1w | 2w | 3w | 4w | 5w | 6w | 7w | 8w | 3 m | 6 m |
| --- | --- | --- | --- | --- | --- | --- | --- | --- | --- | --- | --- |
| **HR +** |  |  |  |  |  |  |  |  |  |  |  |
| TD [mm] | 19.80 | 19.91 | 20.10 | 20.29 | 20.48 | 20.68 | 20.88 | 21.08 | 21.28 | 22.32 | 25.26 |
| 15-yrs mortality rate [%] | 19.57 | 19.73 | 20.02 | 20.31 | 20.61 | 20.91 | 21.21 | 21.51 | 21.82 | 23.4 | 27.8 |
| BC deaths per cohort [n] | 196 | + 2 | + 5 | + 7 | + 10 | + 13 | + 16 | + 19 | + 23 | + 38 | + 82 |
| **HR -** |  |  |  |  |  |  |  |  |  |  |  |
| TD [mm] | 19.80 | 20.06 | 20.51 | 20.98 | 21.45 | 21.94 | 22.44 | 22.95 | 23.47 | 26.26 | 35.17 |
| 15-yrs mortality rate [%] | 19.57 | 19.96 | 20.65 | 21.36 | 22.09 | 22.83 | 23.59 | 24.36 | 25.15 | 29.26 | 40.46 |
| BC deaths per cohort [n] | 196 | + 4 | + 11 | +18 | + 25 | +33 | + 40 | + 48 | + 56 | + 97 | + 209 |

HR, hormone receptor status, m, months, TD, tumor diameter, w, week(s), yrs, years

For both the U.S. and Germany, real-world data on surgical treatment delay are available, and their distributions are shown in Table 2. Frequencies for the first four weeks in the U.S. were split according to data from Germany. The middle of each week represents the duration of delay, for >6/ >12 weeks 48/90 days were assumed for Germany and U.S. It should be added that in Germany 51% of BC patients were treated within 14 days until 2006, this proportion decreased to 17% in 2017 indicating an increase in time between initial diagnosis and start of treatment. In 2020 the number of annual new cases are 287,850/71,375 BCs in the U.S. and Germany, respectively. The exclusion of 6.2% primary M1 cases results in 270,000/66,950 T-N-M0 BCs. These represent the at-risk populations, which can be affected by surgical treatment delay (Table 2). The delay stratified according to HR+/HR- can be associated with an estimated 3,261/1,415 deaths in the U.S. and 640/278 deaths in Germany. Based on these results two relations can be generated:

**Table 2:**
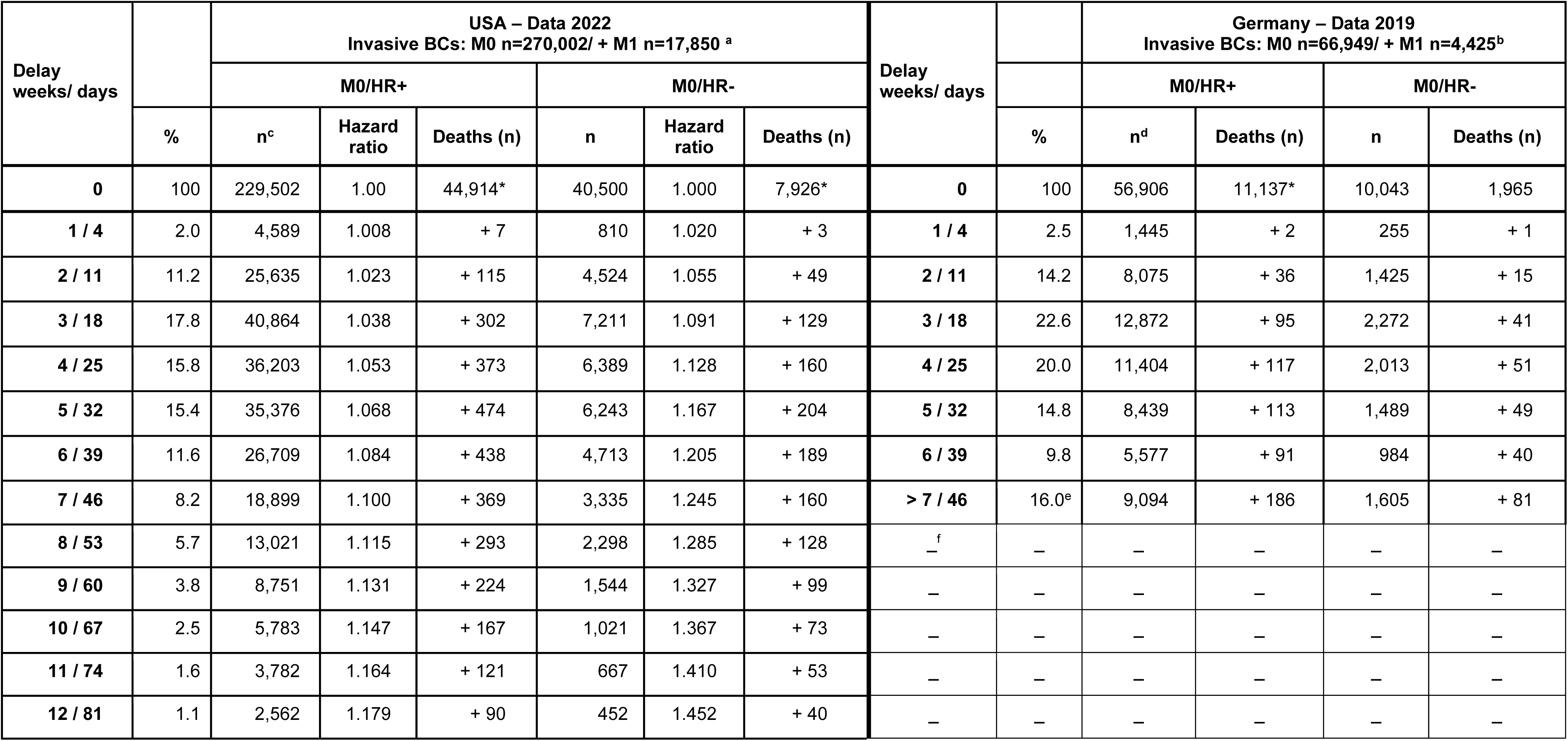
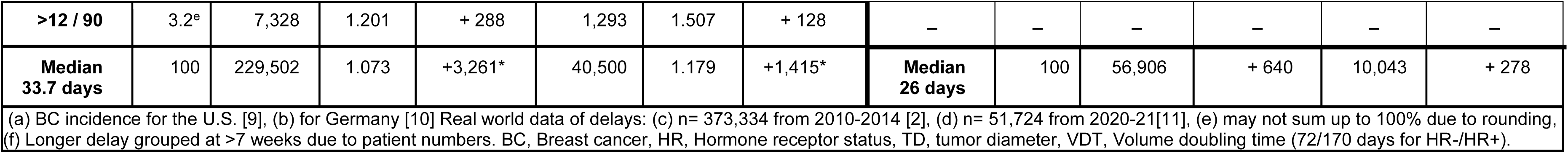
Increase in TD as a function of HR+/HR- and delayed surgery in weeks and mortality caused by delay in the U.S. and Germany. 6.2 % for primary advanced BCs (M1), 85% for HR+ and 19.8 mm TD are assumed. When calculating with a 20.8 mm TD, the values marked with * result higher: In the U.S. with HR+/HR- 48,402/8,541 deaths and 3,429/1,483 deaths by delay, in Germany 12,001/2118 deaths and 675/289 deaths by delay.

Firstly, after 15 years prognosis worsens by 1.6/1.2% for the U.S. and Germany respectively. The difference results from the different distributions of treatment delay with a mean of 33.7 in the U.S. and 26.0 days in Germany.

Secondly, reference to the at-risk populations HR+/ HR- T-N-M0 BCs yields an increase of 7.3/19.7% and 5.7/14.1% BC-associated deaths (U.S./Germany; see Table 2). If all surgical procedures were postponed by 8 weeks, an estimated additional 5,170/2,260 BC deaths would be associated with this delay in the U.S. according to Table 1 for HR+/HR-.

The results are replicated for HR- in a multivariate Cox model with a continuous independent delay variable (delay in days for M0/HR+: 1.000 [95%CI 0.999-1.000], p=0.621; M0/HR-: 1.004 hazard ratio [95%CI 1.002-1.006], p=0.003).

In Table 2, the population is the only varying variable. The other factors such as HR status, increase in TD with delay, association of TD with prognosis, and proportion of M1 remain constant. The mortality caused by the delay can be calculated from available data: Mortality without delay adds up to 70,690/17,527 and with delay to 75,366/18,445 for the U.S. and Germany. Relative to incidence, this results in a 15-year mortality of 26.2/25.8% In 2020 18,425 BC-dependent deaths were registered in Germany.[10] In the U.S., only 43,250 BC deaths are estimated in 2022.[9]

The time-distribution of the deaths caused by surgical treatment delay can be inferred from the distributions of MET-free times (Figure 1D). Since these METs are initiated during the delay interval, the median growth time of MET - twice the above percentiles – and 28 months of median survival from MET result in a median survival times of 12.4/6.5 years of delay-related deaths contribute to mortality up to 30 years which is also suggested by the literature (Figure 1E).[15 16] MCR data through 2010 on HR+ BCs partially suggest this relationship (Figure 1B). Only a few MET still occur 20 years after diagnosis, but can still be the cause of death after 25-30 years. Even 30 years after initial diagnosis new MET are registered in first single tumors. After 25-30 years, patients who were 50.8 (SD ±8.5) years old at diagnosis are on average 77.9 (SD ±8.5) years old. According to our data worse prognostic factors such as a negative HR status or larger TD were not associated with a shorter treatment waiting period (suppl. Table S1).

## Discussion

In this simulation study publicly available, real-world data are used to estimate and compare the risks associated with treatment delay in the U.S. and Germany (Table 2). Based on current data on surgical treatment delay overall prognosis decreases by an estimated 1.6/1.2% for the U.S. and Germany respectively. Depending on HR status postponing surgical treatment by the proposed eight weeks would result in an additional 5,170 HR+/2,260 HR- BC deaths in the U.S. The quantification of the risk associated with a delay in surgical treatment and the underlying MET process requires 30 years of BC survivorship surveillance from cancer registries.

These findings are in line with previous research, for example a study on Medicare BC patients from the U.S. revealed an increasing time interval between the first physician visit and surgery (21 days in 1992 vs. 32 days in 2005).[17] A subsequent assessment of the outcome from 2016 found an increase in all-cause mortality of 9% for each month of delay.[18 19]

The results are based on a comprehensible transformation of valid data and the following applies to both the U.S. and Germany: BC tumor biology, the proportion of M1 at 6.2%, and HR+ status at 85% remain constant. The same may be said regarding current therapy options, these are also available to patients in both countries and should result in similar survival. Long-term survival as a function of HR status is similar after 15 years (Figure 1B).[20], [8] This is also shown by the distributions of MET-free times and the ratio of their medians 60.4/25.2 or 2.40 (Figure 1D) as the ratio 2.36 of VDTs adopted for HR+/HR- of 170/72. The median TD of 19.8 mm used in our simulation study for T-N-M0 BCs remains constant and transferable, because the pT-category distribution is comparable in both populations.[4]

The functional relationship such as the one described above suggests interpolations. Within two weeks, the TD of a HR+ M0 BC of 19.8 mm grows by 0.38 mm and the mortality risk increases mathematically by 0.0058%. However, this risk, which is not relevant in the individual case, would already be implicated in 1,328 additional deaths after 15 years out of 229,502 HR+/M0 BCs, which are diagnosed annually in the U.S. (Table 2). This exemplifies that worldwide a very small risk in a large population can be more severe than a large risk in a small population. Considering screening this means a mortality decrease by 0.8% when TD is diagnosed 0.50 mm earlier.

A risk-free time from diagnosis to treatment start cannot depend on detection, but only on tumor size. For example, if a BC has not initiated MET for eight weeks according to Wiener and was not detected within this interval, it would grow for eight weeks without MET.[2] If it was diagnosed right after this time point, there would be another eight and any number of recurrent weeks without risk of MET. Risk-free interval and MET are mutually exclusive, and MET is a continuous process that follows the principle of “natura non facit saltus”. The time interval after diagnosis and before initial surgical treatment establishes the time of MET initiation and therefore implicates an excess mortality after a median of 6.5/12.4 years. With a 5-year follow-up and 370,000 patients, this association is unlikely to be proven even for triple negative BC.[2] If there is no risk-free delay, it is not possible to specify a limit at which undertreatment begins. It should be noted that a reasonably prioritized treatment of delay-sensitive HR- and larger BCs is not evident in our data (Table S1).

The organization of mammography screening should also be critically reviewed with respect to prolonged time to treatment begin. Prolongation begins with the suspicious finding during mammography, many days before a needle biopsy is performed and a definitive histology is available. If a patient with a suspicious finding is recalled and subsequently diagnosed with an invasive BC, the initial TD on the earlier abnormal mammogram should be calculated retrospectively.[21] This screening time interval represents an additional delay, even before the usual waiting period until initial surgical treatment is performed. Even if screening-detected BCs had a smaller TD, 14.8 mm in our data, delays would predictably enlarge the mortality risk.

Our analyses and concluded thesis carry inherent limitations. One lies in the dependence of the delay effect on the basic distribution of M1, HR-status, and the TD. Especially TD may vary in populations due to screening programs. Because of this dependence and innovations in treatment and diagnosis, the cure rate, which can be estimated by the complement of the relation of mortality and incidence, varies. The official mortality of a year approximately summarizes disease courses which began a few months or up to 30 years ago. Here, the period survival can be used to project disease courses of a year into the future up to a few months up to 30. The calculations for the delay cohorts of Table 2 can be demonstrated with historical cohorts (Fig.1D) but cannot be confirmed experimentally with a randomized trial. However, this does not call into question the dependence on TD and thus the action relevant delay effect.

Table 2 also shows the results using a TD of 20.8 mm instead of 19.8 mm. With a larger TD, the delay effect also increases and exceeds the official mortality rate for Germany by 5.8%. However, we consider a TD close to 20.8mm to be justifiable. First, mortality derived by MCR data is underestimated because the treatment waiting period has increased distinctly since 2016, and this delay-subsequent effect on mortality will appear at the earliest after 10 or more years (i.e. 2026). Second, the BC incidence has increased in the two past decades, therefore the ratio of the mortality to incidence during this time becomes smaller from year to year. Based on these numbers the cure rate is likely underestimated. Furthermore, the neoadjuvant treated BCs are not included in Figure 1A due to missing data on TD at initial diagnosis. For an increase in mean diameter from 19.8 to 20.8 mm, all neoadjuvant treated HR- BCs would have to have a TD of 26.5 mm. In addition, adjuvant therapies are also likely to partially eradicate MET initiated by delay. In addition, by considering 15 instead of 25/30 years of follow-up mortality is underestimated. Assuming a TD of 20.8 mm would result in 74,800/18,540 deaths without delay for the U.S. and Germany. Therefore, a TD value between 19.8 and 20.8 mm may be the best estimate for the two populations.

Interestingly, our simulation results for the U.S. estimate 70,687 BC deaths per year without delay (including M1) and a mortality of 24.6% after 15 years which would appear plausible. However, this is inconsistent with the officially reported 15.1% or 43,250 deaths.[9 22 23] The 10-year relative survival with 84.7% already results in 44,041 deaths.[24] With the modeled rate of 86.2%, the 43,250 deaths will already be reached after 11.9 years.[25] Our estimates of 25-year survival from SEER data of 69,660 deaths are almost equivalent to the expected mortality of 70,687 deaths without delay plus 4,676 delay-associated deaths. The reason for this inconsistency remains unclear and should be examined further.

Estimates of the delay effect show that every day counts until initial surgery. The delay time can be important both for the health care system and the patient. On the one hand there are possible sociodemographic and socioeconomic factors, preoperative diagnostic procedures such as imaging and histopathological examination and a comprehensive examination of the patient to determine which treatment options can be offered, their prospects of success, and an assessment of the patient’s physical and psychological condition.[17] From the patient’s point of view, this time is important to come to terms with the diagnosis and treatments, perhaps with a second opinion, and to mobilize coping strategies.[26]

However, in each individual case, unnecessary delays should be avoided without exerting special pressure concerning the risk of mortality. If prolonged times between first diagnosis and surgery do not serve to improve treatment overall but emerge as bottlenecks, prioritization by HR and TD should be considered and undertaken. Alternatively, neoadjuvant endocrine therapies in particular could be administered.[27] Since the treatment sequences a) neoadjuvant chemotherapy → surgery and b) surgery → adjuvant chemotherapy are equivalent in regard to effect on MET initiation, the consequences of necessary delays can be reduced by prioritizing a neoadjuvant approach. It would reduce time pressure for scheduling surgery and present an advantage with reference to strength of response and TD reduction.

It can be concluded that there is no risk-free time from diagnosis to treatment. A growing tumor can continuously initiate MET, and therefore pose a relevant risk factor impacting prognosis with every additional millimeter. Confirmed BCs should be treated according to HR status as soon as possible. A similar association may be assumed for any invasive cancer. Therefore, delays in treatment present a problem in healthcare systems worldwide.

## Data availability statement

The datasets used to support the results of this study are publicly available (SEER, RKI, publications). Data from the MCR are protected by data privacy protection jurisdiction but may be made available in anonymized form on reasonable request via the corresponding author: KH.

## Author Contributions

Concept and design: Hölzel, Halfter

Acquisition, analysis, interpretation of data: All authors Drafting of the manuscript: Hölzel, Schlesinger-Raab, Halfter

Critical revision of the manuscript for intellectual content: Schubert-Fritschle Statistical analysis: Hölzel, Halfter, Schlesinger-Raab

Administration, technical, material support: Schubert-Fritschle Supervision: Hölzel, Schubert-Fritschle

## Funding

Not applicable

## Conflict of Interest Disclosures

The authors declare no conflict of interest to disclose.

## Acknowledgment

We would like to thank all former MCR employees for their contributions to the excellent data quality.

## Notes

### Competing Interest Statement

The authors have declared no competing interest.

### Funding Statement

This study did not receive any funding

### Author Declarations

The study used ONLY openly available human data that were originally located at: SEER: https://seer.cancer.gov/statistics-network/ RKI: https://www.krebsdaten.de/Krebs/SiteGlobals/Forms/Datenbankabfrage/datenbankabfrage_stufe2_form.html MCR: http://www.tumorregister-muenchen.de/en/facts/specific_analysis.php

