## Supplemental Table 1 for "Prolonged time to breast cancer surgery and the risk for metastasis"

Table of Contents

Table S1 Pages 3-4

**Table S1:** Cohort of first single primary M0 invasive breast cancer 1998-2010 stratified by delay between initial diagnosis and treatment

| **Time of delay in days** | | **n.a. / 0 days** | | **1-14 days** | | **15-35 days** | | **> 35 days** | | **Total** | | **p-value** |
|---|---|---|---|---|---|---|---|---|---|---|---|---|
| | | **n=7062** | | **n=10562** | | **n=5693** | | **n=1135** | | **n=24452** | | |
| **Prognostic factors** | | **n** | **(%) [a]** | **n** | **(%) [a]** | **n** | **(%) [a]** | **n** | **(%) [a]** | **n** | **(%) [a]** | |
| **Age** | mean ± SD | 61.5 | ± 13.7 | 61.2 | ± 13.2 | 60.8 | ± 12.5 | 61.7 | ± 14.2 | 61.2 | ± 13.2 | 0.0798 n.s. |
| **Age in classes** | < 50 yrs. | 1532 | (21.7) | 2368 | (22.4) | 1217 | (21.4) | 254 | (22.4) | 5371 | (22.0) | <0.0001 |
| | 50-59 yrs. | 1711 | (24.2) | 2415 | (22.9) | 1379 | (24.2) | 253 | (22.3) | 5758 | (23.6) | |
| | 60-69 yrs. | 1913 | (27.1) | 3056 | (28.9) | 1843 | (32.4) | 330 | (29.1) | 7142 | (29.2) | |
| | 70-79 yrs. | 1197 | (17.0) | 1826 | (17.3) | 890 | (15.6) | 168 | (14.8) | 4081 | (16.7) | |
| | ≥ 80 yrs. | 709 | (10.0) | 897 | (8.5) | 364 | (6.4) | 130 | (11.5) | 2100 | (8.6) | |
| **c/p T-Category** | T1 | 4230 | (59.9) | 5984 | (56.7) | 3420 | (60.1) | 612 | (53.9) | 14246 | (58.3) | <0.0001 |
| | T2 | 2180 | (30.9) | 3771 | (35.7) | 1782 | (31.3) | 371 | (32.7) | 8104 | (33.1) | |
| | T3 | 253 | (3.6) | 401 | (3.8) | 255 | (4.5) | 73 | (6.4) | 982 | (4.0) | |
| | T4 | 267 | (3.8) | 363 | (3.4) | 177 | (3.1) | 62 | (5.5) | 869 | (3.6) | |
| | n.a. | 132 | (1.9) | 43 | (0.4) | 59 | (1.0) | 17 | (1.5) | 251 | (1.0) | |
| **c/p N-Category** | N0 | 4137 | (58.6) | 6399 | (60.6) | 3657 | (64.2) | 673 | (59.3) | 14866 | (60.8) | <0.0001 |
| | N+ | 2256 | (32.0) | 3789 | (35.9) | 1897 | (33.3) | 390 | (34.4) | 8332 | (34.1) | |
| | NX | 527 | (7.5) | 355 | (3.4) | 130 | (2.3) | 68 | (6.0) | 1080 | (4.4) | |
| | n.a. | 142 | (2.0) | 19 | (0.2) | 9 | (0.2) | 4 | (0.4) | 174 | (0.7) | |
| **Grade** | G1 | 954 | (13.5) | 1305 | (12.4) | 822 | (14.4) | 160 | (14.1) | 3241 | (13.3) | <0.0001 |
| | G2 | 3876 | (54.9) | 6099 | (57.7) | 3388 | (59.5) | 655 | (57.7) | 14018 | (57.3) | |
| | G3 | 2062 | (29.2) | 3049 | (28.9) | 1442 | (25.3) | 310 | (27.3) | 6863 | (28.1) | |

| | | | | | | | | | | | | |
|---|---|---|---|---|---|---|---|---|---|---|---|---|
| | n.a. | 170 | (2.4) | 109 | (1.0) | 41 | (0.7) | 10 | (0.9) | 330 | (1.4) | |
| **Hormone receptor status** | HR+ | 5747 | (81.4) | 9161 | (86.7) | 4985 | (87.6) | 966 | (85.1) | 20859 | (85.3) | <0.0001 |
| | HR- | 778 | (11.0) | 1236 | (11.7) | 635 | (11.2) | 145 | (12.8) | 2794 | (11.4) | |
| | n.a. | 537 | (7.6) | 165 | (1.6) | 73 | (1.3) | 24 | (2.1) | 799 | (3.3) | |
| **HER2 status** | HER2+ | 645 | (9.1) | 1205 | (11.4) | 684 | (12.0) | 150 | (13.2) | 2684 | (11.0) | <0.0001 |
| | HER2- | 3433 | (48.6) | 7399 | (70.1) | 4427 | (77.8) | 823 | (72.5) | 16082 | (65.8) | |
| | HER2 unclear | 234 | (3.3) | 363 | (3.4) | 139 | (2.4) | 32 | (2.8) | 768 | (3.1) | |
| | n.a. | 2750 | (38.9) | 1595 | (15.1) | 443 | (7.8) | 130 | (11.5) | 4918 | (20.1) | |
| **Subtype** | Luminal-A like | 1463 | (20.7) | 4567 | (43.2) | 3054 | (53.6) | 547 | (48.2) | 9631 | (39.4) | <0.0001 |
| | Luminal-B like HER2- | 363 | (5.1) | 1241 | (11.8) | 793 | (13.9) | 139 | (12.3) | 2536 | (10.4) | |
| | Lumnal-B like HER2+ | 253 | (3.6) | 725 | (6.9) | 444 | (7.8) | 90 | (7.9) | 1512 | (6.2) | |
| | HER2- non-luminal | 94 | (1.3) | 261 | (2.5) | 170 | (3.0) | 40 | (3.5) | 565 | (2.3) | |
| | Triple negative | 202 | (2.9) | 600 | (5.7) | 352 | (6.2) | 79 | (7.0) | 1233 | (5.0) | |
| | n.a. | 4687 | (66.4) | 3168 | (30.0) | 880 | (15.5) | 240 | (21.2) | 8975 | (36.7) | |
| **Surgery** | Breast conserving | 4929 | (69.8) | 8095 | (76.6) | 4259 | (74.8) | 765 | (67.4) | 18048 | (73.8) | <0.0001 |
| | Mastectomy | 1747 | (24.7) | 2394 | (22.7) | 1402 | (24.6) | 359 | (31.6) | 5902 | (24.1) | |
| | n.a. | 386 | (5.5) | 73 | (0.7) | 32 | (0.6) | 11 | (1.0) | 502 | (2.0) | |
| **Neoadjuvant** systemic treatment | yes | 343 | (4.9) | 278 | (2.6) | 419 | (7.4) | 139 | (12.3) | 1179 | (4.8) | <0.0001 |
| | no | 6719 | (95.1) | 10284 | (97.4) | 5274 | (92.6) | 996 | (87.8) | 23273 | (95.2) | |

SD: standard deviation, n.s.: not significant as defined by a level α set at 0.05, n.a. not available

[a] Missing values were not excluded from calculations of frequency distribution, column percentage can differ slightly from 100% due to rounding.
